# Moderate-to-vigorous physical activity, longitudinal blood pressure trajectories, and hypertension-related outcomes in older adults at high cardiovascular risk: a cohort study

**DOI:** 10.64898/2026.11.17.26368306

**Authors:** Eleonora Fornara, Marta H. Hernández, Alberto Goday, Anna Camps-Vilaró, Estefanía Toledo, Jordi Salas-Salvadó, Ramón Estruch, Emilio Ros, Raquel Cueto-Galan, Miquel Fiol, José Lapetra, Juan Timiraos, Lluís Serra-Majem, Xavier Pintó, Miguel Ruiz-Canela, Héctor Vázquez-Lorente, Montserrat Fitó, Álvaro Hernáez

## Abstract

**Background:** We aimed to investigate the association between moderate-to-vigorous leisure-time physical activity (MVLTPA) and longitudinal trajectories of systolic (SBP) and diastolic (DBP) blood pressure, as well as the incidence of high blood pressure and antihypertensive drug initiation, in adults aged 60-80 years at high cardiovascular risk.

**Methods:** In participants from the PREDIMED trial (61% women; 5,254 participants), MVLTPA was assessed using the Minnesota Leisure Time Physical Activity Questionnaire and expressed as cumulative average metabolic equivalent of task·minutes/day. SBP and DBP trajectories were modelled using mixed-effects models with cubic splines (≥100 [adequate] vs <100 [insufficient] metabolic equivalent of task·minutes/day). First occurrence of high blood pressure (SBP ≥140 mmHg or DBP ≥90 mmHg) and antihypertensive drug initiation were assessed with Cox models (adequate vs insufficient, and continuous MVLTPA).

**Results:** SBP and DBP declined with increasing age, with no clinically meaningful differences between MVLTPA groups. The association between adequate MVLTPA and incident high blood pressure was null (HR 1.11, 95% CI 0.94-1.32). However, adequate MVLTPA was associated with a 17% lower risk of antihypertensive drug initiation (HR 0.83, 95% CI 0.71-0.98). The association appeared stronger in women (HR 0.74, 95% CI 0.58-0.94) than in men (HR 0.98, 95% CI 0.78-1.22) (*p*-value for sex interaction = 0.058). A modest inverse linear dose-response association between MVLTPA and risk of antihypertensive drug initiation was also observed (per 25 metabolic equivalent of task·minutes/day: HR 0.99, 95% CI 0.97-1.00; *p*-value = 0.022).

**Conclusions:** In older adults at high cardiovascular risk, MVLTPA was not associated with clinically meaningful differences in SBP or DBP trajectories or incident high blood pressure, but was associated with a lower risk of antihypertensive drug initiation. These findings support promoting habitual physical activity as part of cardiovascular risk management in older adults.

**Trial registration:** ISRCTN35739639

## INTRODUCTION

Hypertension, defined as systolic blood pressure (SBP) ≥140 mmHg or diastolic blood pressure (DBP) ≥90 mmHg [1] is the leading preventable cause of premature death worldwide. It affects an estimated 1.4 billion people globally and significantly raises the risk of coronary heart disease, stroke, and kidney disease [2], often requiring pharmacological treatment when lifestyle measures alone are insufficient. Among such lifestyle measures, leisure-time physical activity (LTPA) is consistently recommended for both the prevention and management of hypertension [3]. In adults and older adults, current European guidelines recommend at least 150 min/week of moderate-intensity exercise or 75 min/week of vigorous-intensity exercise, complemented by dynamic or isometric resistance training 2-3 times per week [4]. These recommendations are supported by evidence showing that regular supervised training lowers blood pressure by approximately 4.5 mmHg for SBP and 2.5 mmHg for DBP [5], with larger reductions of 5.4 mmHg for SBP and 4.8 mmHg for DBP among individuals with hypertension [6]. Meta-analyses of cohort studies also suggest an association between higher LTPA and a lower risk of incident hypertension [7]. Additionally, cohort studies have linked higher LTPA to a lower risk of antihypertensive drug initiation, with the lowest risk around 180-360 metabolic equivalents of task-minutes per day (MET·min/day) [8].

Despite this evidence, important gaps remain regarding the long-term associations between moderate-to-vigorous LTPA (MVLTPA), the intensity of physical activity emphasized in current recommendations, and blood pressure in older adults. In particular, few studies have examined age-dependent blood pressure trajectories and the incidence of hypertension-related outcomes in this population, despite advancing age being a major risk factor for hypertension [9]. Furthermore, recommendations remain largely uniform across age groups and sexes, although evidence on potential differences in these associations by age and sex is limited [10]. Additional uncertainties concern the associations of MVLTPA with blood pressure among individuals with established hypertension, its dose-response relationship with incident high blood pressure, and potential sex differences in the initiation of antihypertensive medication. Therefore, this study aimed to investigate the association between MVLTPA and blood pressure in older adults at high cardiovascular risk by characterising age-related SBP and DBP trajectories between ages 60 and 80 years, examining the incidence of high blood pressure and antihypertensive drug initiation, and assessing dose-response associations and potential differences by sex.

## METHODS

### Study population

The present study was conducted using PREDIMED data as a prospective cohort. The PREDIMED study was a multicenter, randomized, controlled intervention trial conducted in Spain between 2003 and 2010, aimed at assessing the long-term effects of the Mediterranean diet on the primary prevention of major cardiovascular events in older adults [11]. Eligible participants were men aged 55-80 years and women aged 60-80 years, free of cardiovascular disease at enrolment but presenting type 2 diabetes or at least three major cardiovascular risk factors, including smoking, hypertension, dyslipidaemia, overweight or obesity, and family history of premature coronary heart disease. In our analyses, the study population was restricted to participants aged 60-80 at baseline. As shown in **Figure 1**, participants were excluded from the analyses if they were younger than 60 years at baseline, had missing baseline data on MVLTPA, or had blood pressure measurements available only at baseline. For survival analyses, only participants with the corresponding outcome at baseline were excluded. This study was conducted and reported in accordance with the Strengthening the Reporting of Observational Studies in Epidemiology (STROBE) guidelines.

**Figure 1.**
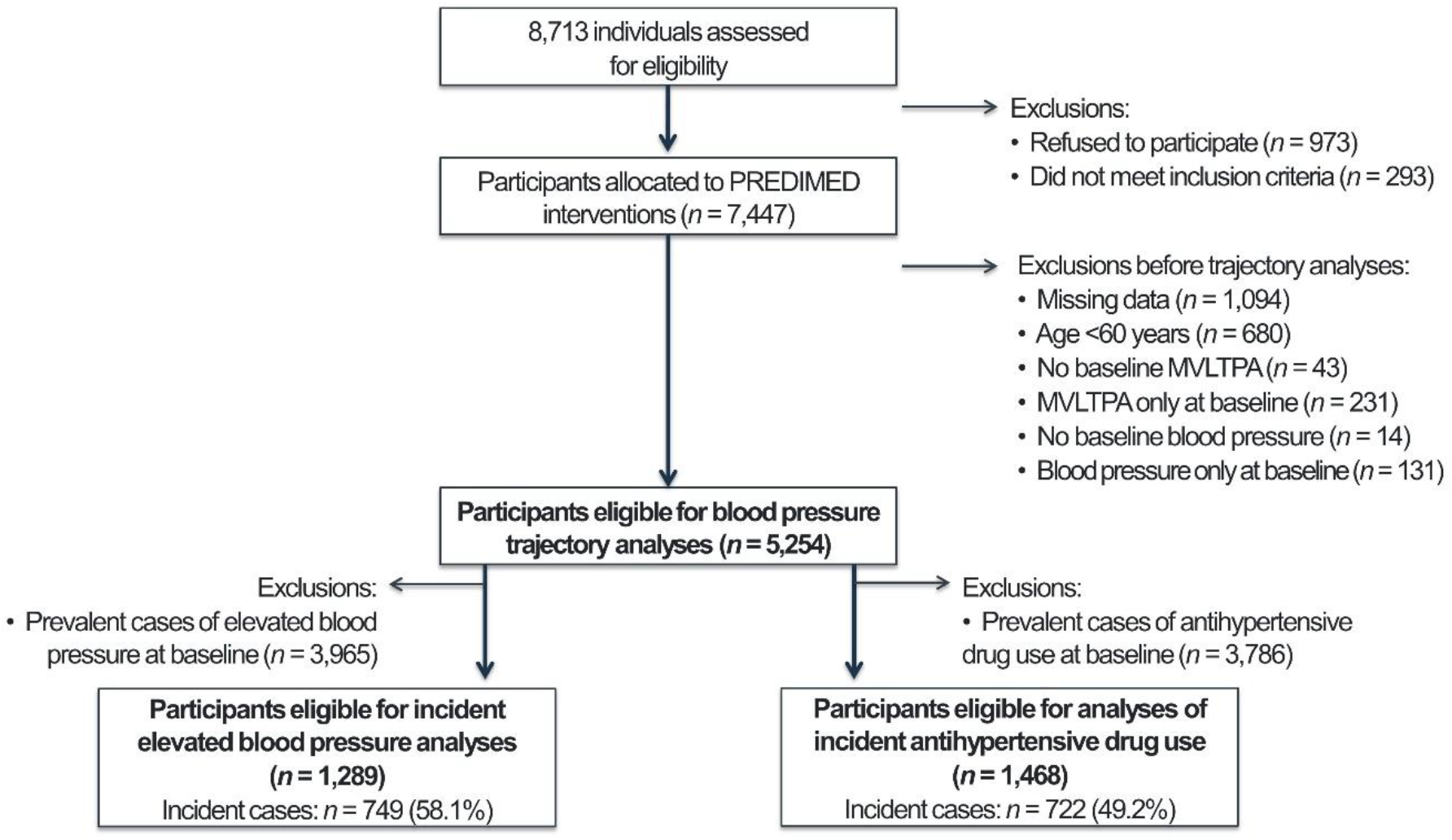
Flow chart.

### Exposure: usual moderate-to-vigorous leisure-time physical activity levels

MVLTPA was estimated using the Minnesota Leisure Time Physical Activity Questionnaire, a tool previously validated in Spanish adult populations of men and women [12,13]. Participants reported the frequency (days) and duration (minutes per day) of 67 activities performed during the previous year. MVLTPA was quantified using MET·min/day and calculated by multiplying the MET value assigned to each activity of moderate and vigorous intensity by its average daily duration and summing across all reported activities. Because MVLTPA was recorded at each study visit, we derived a participant-specific cumulative average from all available measurements, which was in turn dichotomized into adequate (≥100 MET·min/day) and insufficient (<100 MET·min/day, reference) categories [4].

### Outcomes

Trained clinical staff measured systolic and diastolic blood pressure at baseline and at each annual follow-up visit between 2003 and 2010, using a validated semi-automatic device (Omron HEM-705CP, Hoofddorp, the Netherlands). Three readings were taken on the same arm, two minutes apart, with participants seated and at rest for five minutes beforehand; cuff size was selected according to arm circumference, and measurements were taken at heart level. The mean of the second and third readings was recorded as the participant’s blood pressure value for that visit [14]. Data on antihypertensive drug use (yes/no) were likewise collected at baseline and at each yearly visit, covering renin-angiotensin-aldosterone system inhibitors, calcium antagonists, thiazide/thiazide-like/loop diuretics, mineralocorticoid receptor antagonists, beta-blockers, alpha-blockers, and other minor antihypertensive drug families as previously described [8].

With these data, we constructed an age-indexed dataset in which each SBP and DBP measurement was assigned to the participant’s chronological age at the time of assessment [15], so that individuals contributed data only for the ages actually covered by their own follow-up. Age-related trajectories of SBP and DBP between 60 and 80 years were then estimated by pooling, at each age, the records from all participants observed at that age, regardless of whether any single participant had been followed continuously across the entire interval. **Additional File-Supplementary Table 1** reports, by age, the number of SBP and DBP measurements underlying these estimates.

We defined two endpoints: the first occurrence of high blood pressure (SBP ≥140 mmHg or DBP ≥90 mmHg) [1] and initiation of antihypertensive medication. For each endpoint, only participants free of the respective outcome at baseline were considered at risk [8].

### Other variables

In the baseline PREDIMED visit, trained clinical staff collected information on age (years), self-reported sex (women/men), recruitment center, educational level (primary, secondary, higher), smoking status (current, former, or never smoker), energy intake (kcal/day, from a validated 137-item food frequency questionnaire), adherence to a Mediterranean diet (using the 14-point Mediterranean Diet adherence short screener by Schroder H et al.) [16], and prevalence of diabetes (yes/no), hypertension (yes/no), and hypercholesterolemia (yes/no). Participants’ randomized PREDIMED dietary intervention group (Mediterranean diet supplemented with extra-virgin olive oil, Mediterranean diet supplemented with nuts, or low-fat control diet) was also recorded and included as a covariate. Before the analyses, covariates with non-structural missingness at baseline were imputed using additive regression, bootstrapping, and predictive mean matching based on all available information from the other study variables (1.8% for educational level, 1.0% for energy intake, and 0.7% for adherence to the Mediterranean diet).

### Statistical analyses

Characteristics of the participants in the baseline PREDIMED visit were summarized using means and standard deviations for normally distributed continuous variables, medians and interquartile ranges for non-normally distributed variables, and proportions for categorical variables.

### Longitudinal trajectory analyses

We modelled age-dependent trajectories of SBP and DBP, defined as model-predicted mean values at each age between 60 and 80 years, in the pooled longitudinal dataset, using mixed-effects regression models fitted with multivariable smoothed cubic splines (K+4 degrees of freedom) [15,17,18]. An age × MVLTPA category interaction term was included in each model, allowing the shape of the trajectory to vary between the adequate and insufficient MVLTPA groups. Models were adjusted for sex, educational level, PREDIMED intervention group, diabetes, hypertension, hypercholesterolemia, body mass index, smoking status, adherence to a Mediterranean diet, and energy intake. We graphically represented the mean age-related trajectories of SBP and DBP according to MVLTPA category using predicted values, and estimated between-group differences at ages 60, 65, 70, 75, and 80 years using linear regression models. SBP and DBP trajectories were also estimated separately in women and men (after assessing whether trajectories differed by sex using likelihood ratio tests for interaction), and in participants with hypertension at baseline.

### Incident outcome analyses

We examined the association between MVLTPA and the risk of incident high blood pressure and antihypertensive drug initiation using two complementary approaches. In the first, cumulative incidence of each outcome between ages 60 and 80 years was estimated across MVLTPA categories, with 95% confidence intervals, using Kaplan-Meier curves; age served as the underlying time scale, with participants entering at their baseline age and exiting at the age of event occurrence or the age on December 1, 2010, whichever came first. Kaplan-Meier curves were generated in the full study population and separately for women and men, after formally testing whether survival models differed by sex using likelihood ratio tests for interaction. Inverse probability weights, derived from a propensity score model adjusted for the same covariates as the trajectory analyses, were applied throughout to minimize confounding. In the second approach, we used age-as-time-scale multivariable Cox proportional hazards models, adjusted for the same set of covariates, to quantify the association between the cumulative average of MVLTPA and the risk of each outcome. We first estimated hazard ratios (HRs) comparing adequate MVLTPA (≥100 MET·min/day) with insufficient MVLTPA (<100 MET·min/day, reference); then, we modelled MVLTPA as a continuous variable to characterize dose-response associations, using participants reporting zero MET·min/day as the reference group. In these analyses, non-linearity was assessed by comparing a model including only the linear MVLTPA term with a model additionally including smoothed cubic spline terms, using likelihood ratio tests. When evidence of non-linearity was detected, spline-based Cox models were used to characterize the dose-response association and to identify the MVLTPA value associated with the lowest risk of each outcome; segment-specific estimates were reported for ranges showing the strongest risk gradient. For graphical purposes, MVLTPA values were truncated at 500 MET·min/day. As for the categorical analyses, we tested whether the dose-response associations differed by sex using likelihood ratio tests for interaction and presented them separately for women and men accordingly.

All analyses were conducted using R software (version 4.3.1). Code for data management and statistical analyses is available at https://github.com/alvarohernaez/MVLTPA_cardiometab_PREDIMED

### Use of generative artificial intelligence

ChatGPT (OpenAI) was used to assist with language editing, manuscript organization, and refinement of scientific writing. All AI-assisted content was critically reviewed and revised by the authors, who take full responsibility for the accuracy, integrity, and final content of the manuscript.

## RESULTS

### Study participants

The study population consisted predominantly of older adults with a high burden of cardiovascular risk factors. The analyses included 5,254 participants (60% with insufficient and 40% with adequate cumulative average MVLTPA), whose baseline characteristics by MVLTPA category are shown in **Table 1**. Participants with adequate MVLTPA consistently exhibited a more favourable sociodemographic and cardiometabolic profile, including a lower prevalence of hypertension, obesity, central adiposity, and hypertriglyceridemia, as well as greater adherence to the Mediterranean diet, compared with those with insufficient MVLTPA.

**Table 1.** Characteristics of the study participants in the PREDIMED baseline visit.

|  | All participants<br>(*n* = 5,254) | Insufficient MVLTPA<br>(*n* = 3,124) | Adequate MVLTPA<br>(*n* = 2,130) | *p*-value |
|---|---|---|---|---|
| Age, years (mean ± SD) | 68.2 ± 5.4 | 68.9 ± 5.6 | 67.1 ± 5.0 | <0.001 |
| Females, *n* (%) | 3,185 (60.6%) | 2,199 (70.4%) | 986 (46.3%) | <0.001 |
| Secondary/higher education, *n* (%) | 1,085 (20.7%) | 565 (18.1%) | 520 (24.4%) | <0.001 |
| Type-II diabetes, *n* (%) | 2,600 (49.5%) | 1,552 (49.7%) | 1,048 (49.2%) | 0.734 |
| Hypertension, *n* (%) | 4,364 (83.1%) | 2,645 (84.7%) | 1,719 (80.7%) | <0.001 |
| Hypertriglyceridemia, *n* (%) | 1,498 (28.5%) | 940 (30.1%) | 558 (26.2%) | 0.002 |
| Hypercholesterolemia, *n* (%) | 3,749 (71.4%) | 2,224 (71.2%) | 1,525 (71.6%) | 0.807 |
| Smoking habit: |  |  |  | <0.001 |
| Never smokers, *n* (%) | 3,356 (63.9%) | 2,159 (69.1%) | 1,197 (56.2%) |  |
| Former smokers, *n* (%) | 644 (12.3%) | 351 (11.2%) | 293 (13.7%) |  |
| Current smokers, *n* (%) | 1,254 (23.9%) | 613 (19.6%) | 641 (30.1%) |  |
| Body mass index categories: |  |  |  | <0.001 |
| <25.0 kg/m<sup>2</sup>, *n* (%) | 391 (7.44%) | 207 (6.63%) | 184 (8.63%) |  |
| 25.0-29.9 kg/m<sup>2</sup>, *n* (%) | 2,425 (46.2%) | 1,274 (40.8%) | 1,151 (54.0%) |  |
| ≥30.0 kg/m<sup>2</sup>, *n* (%) | 2,438 (46.4%) | 1,642 (52.6%) | 796 (37.4%) |  |
| Adherence to a Mediterranean diet (mean ± SD) | 8.7 ± 1.9 | 8.6 ± 1.9 | 8.9 ± 1.8 | <0.001 |
| Energy intake, kcal/day<br>(mean ± SD) | 2,261 ± 581 | 2,224 ± 579 | 2,316 ± 581 | <0.001 |

### Trajectories of systolic and diastolic blood pressure

SBP tended to decline with increasing age in all participants, with slightly lower values in individuals with adequate MVLTPA (although between-group differences were not clinically meaningful), which were more apparent when women and men were studied separately (women: differences ranging from −1.7 to +1.0 mmHg; men: differences ranging from −3.0 to +0.4 mmHg) (**Figure 2**; exact between-group differences are shown in **Additional File-Supplementary Figure 1**).

**Figure 2.**
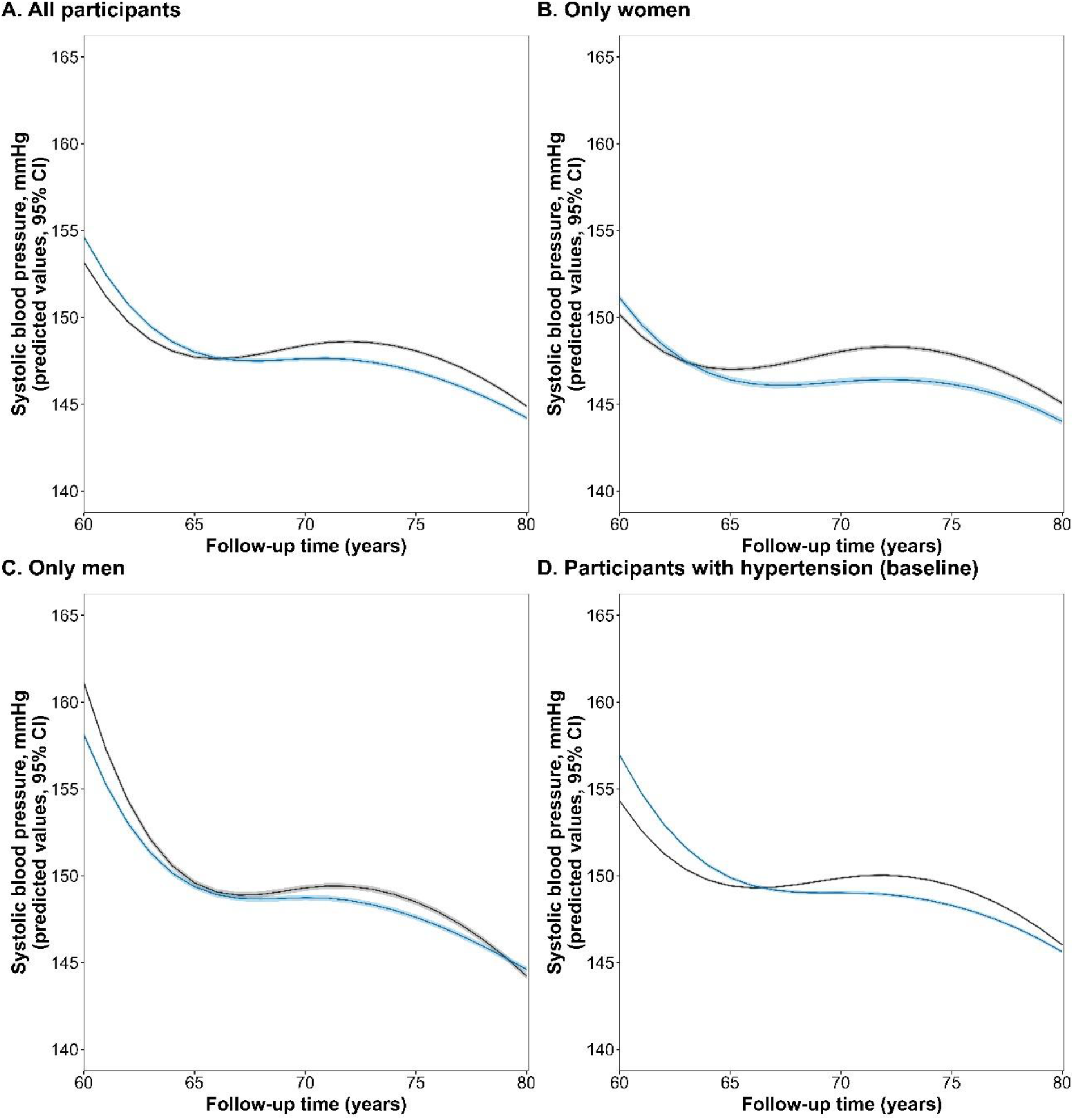
Age-related trajectories of SBP according to cumulative average MVLTPA category. Panels show trajectories (with 95% CI) of predicted mean SBP in all participants (A), women (B), men (C), and participants with hypertension at baseline (D). Blue represents participants with adequate MVLTPA (≥100 MET·min/day); grey represents participants with insufficient MVLTPA (<100 MET·min/day).

DBP also declined progressively with increasing age in all participants, with non-clinically meaningful between-group differences between individuals with adequate and insufficient MVLTPA (women: differences ranging from −0.7 to + 1.6 mmHg; men: differences ranging from −1.6 to +1.8 mmHg). (**Figure 3**; exact between-group differences are available in **Additional File-Supplementary Figure 1B**).

**Figure 3.**
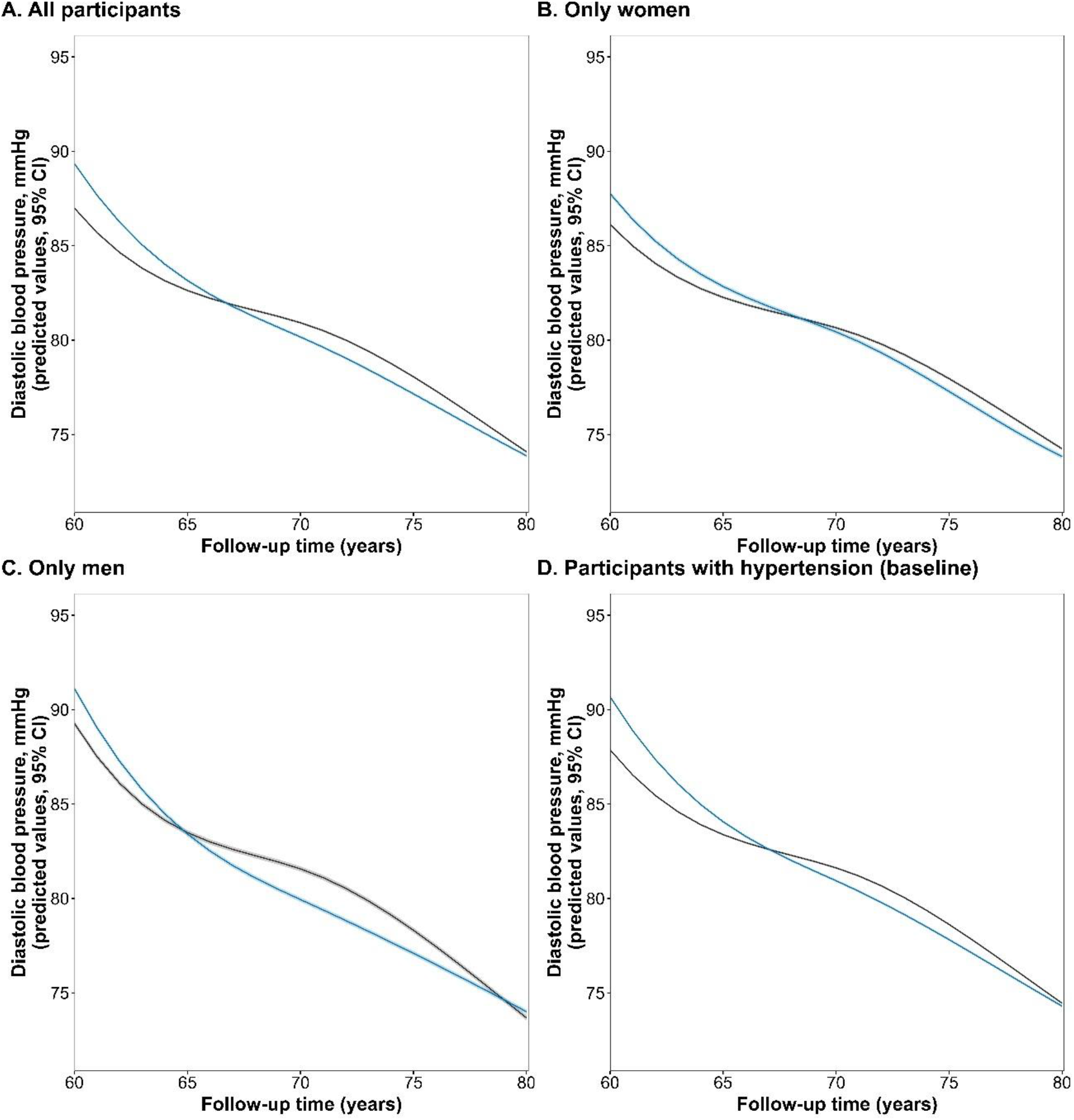
Age-related trajectories of DBP according to cumulative average MVLTPA category. Panels show trajectories (with 95% CIs) of predicted mean DBP in all participants (A), women (B), men (C), and participants with hypertension at baseline (D). Blue represents participants with adequate MVLTPA (≥100 MET·min/day); grey represents participants with insufficient MVLTPA (<100 MET·min/day).

### Onset of high blood pressure and antihypertensive drug initiation

Among participants without high blood pressure at the baseline PREDIMED visit (*n* = 1,289), incidence of high blood pressure was observed in 58.1% of participants between 60 and 80 years of age. Adequate MVLTPA was not significantly associated with incident high blood pressure in the total study population (HR 1.11, 95% CI 0.94 to 1.32). Point estimates were similar between women (HR 1.10, 95% CI 0.88 to 1.37) and men (HR 1.09, 95% CI 0.83 to 1.44), with no evidence of a sex interaction (*p*-value for interaction = 0.895; **Figure 4B-C**). Dose-response analyses did not support a significant linear or non-linear association between MVLTPA and incident high blood pressure (**Figure 4D**), either in women or in men (*p*-value for interaction = 0.553; **Figure 4E**).

**Figure 4.**
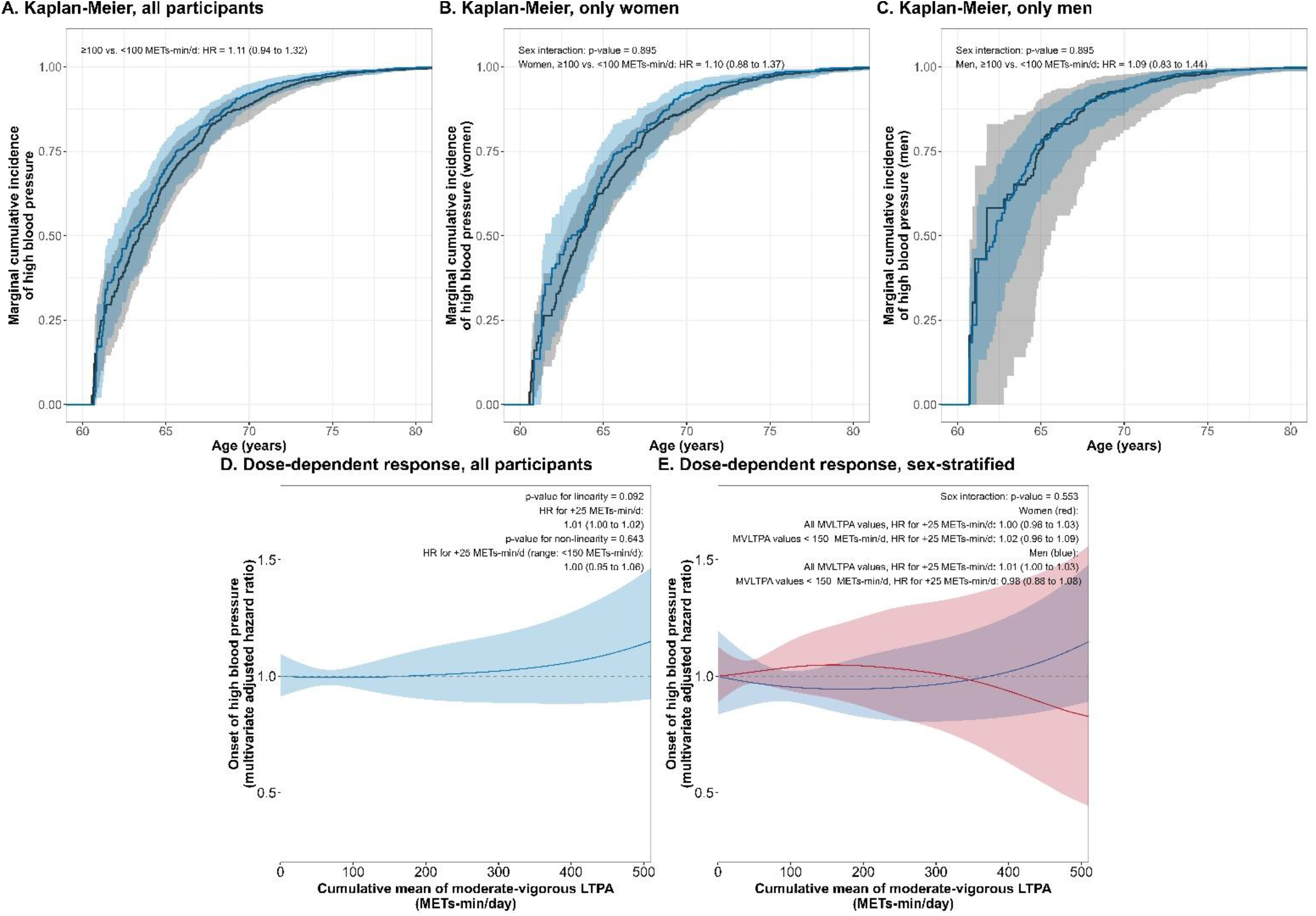
Association between cumulative average MVLTPA and incident high blood pressure between 60-80 years. Panels show Kaplan-Meier curves in all participants (A), women (B), and men (C); the dose-response association in the overall population (D), and sex-specific dose-response associations (E). In panels A-C, blue represents participants with adequate MVLTPA (≥100 MET·min/day) and grey represents participants with insufficient MVLTPA (<100 MET·min/day). In panel E, red represents women and blue represents men.

Among participants without antihypertensive treatment at the baseline PREDIMED visit (*n* = 1,468), incidence of antihypertensive drug initiation was observed in 49.2% of participants between 60 and 80 years of age. Adequate MVLTPA was associated with a 17% lower risk of antihypertensive drug initiation (HR 0.83, 95% CI 0.71 to 0.98). An interaction by sex was suggested (*p*-value for interaction = 0.058), and the association appeared stronger in women (HR 0.74, 95% CI 0.58 to 0.94) than in men (HR 0.98, 95% CI 0.78 to 1.22; **Figure 5B-C**). The dose-response analyses suggested a linear association between greater MVLTPA and lower risk of initiation of antihypertensive medication (per +25 METs-min/d, HR 0.99, 95% CI 0.97 to 1.00, *p*-value = 0.022; *p*-value for non-linearity = 0.222; **Figure 5D**). There was no evidence of an interaction by sex in the dose-response analyses (*p*-value for sex interaction = 0.243; **Figure 5E**).

**Figure 5.**
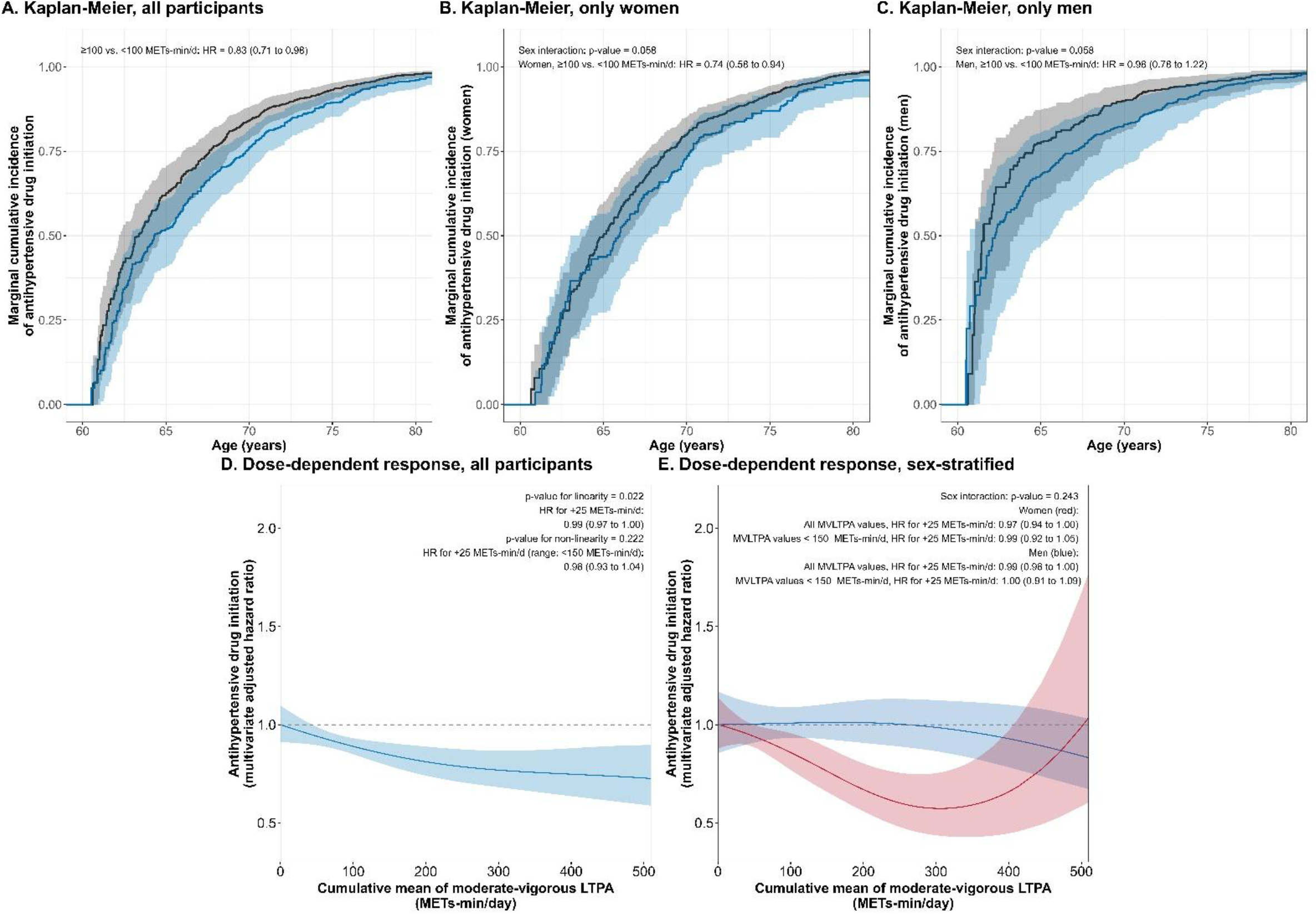
Association between cumulative average MVLTPA and antihypertensive drug initiation between 60-80 years. Panels show Kaplan-Meier curves in all participants (A), women (B), and men (C); the dose-response association in the overall population (D), and sex-specific dose-response associations (E). In panels A-C, blue represents participants with adequate MVLTPA (≥100 MET·min/day) and grey represents participants with insufficient MVLTPA (<100 MET·min/day). In panel E, red represents women and blue represents men.

## DISCUSSION

In this prospective cohort of older adults at high cardiovascular risk, both SBP and DBP tended to decline with increasing age, with no clear or clinically meaningful differences between participants with adequate and insufficient MVLTPA. Consistently, adequate MVLTPA was not associated with a lower risk of incident high blood pressure.

However, adequate MVLTPA was associated with a 17% lower risk of antihypertensive drug initiation, with a stronger association observed in women than in men. Dose-response analyses further suggested a modest linear association between greater MVLTPA and a lower risk of antihypertensive drug initiation.

Our findings contrast with previous meta-analyses showing that exercise training modestly reduces SBP (by 4-5 mmHg) and DBP (by 3-5 mmHg) [5,6] and the inverse association between LTPA and hypertension risk [7]. Several factors may explain these differences. First, previous studies assessed structured exercise interventions, whereas we examined habitual MVLTPA in older adults at high cardiovascular risk, most of whom already had hypertension. Habitual MVLTPA may produce smaller blood pressure reductions than supervised exercise, consistent with evidence only showing 3 mmHg greater SBP reductions with supervised exercise but no significant between-group differences in DBP [19,20]. Second, the high burden of cardiovascular risk factors and widespread antihypertensive treatment in an older adult population may have obscured modest differences between activity groups [21].

Notably, 83.1% of participants already had hypertension at baseline, limiting the population at risk of incident high blood pressure and potentially reducing our ability to detect an association with MVLTPA. Third, the age-related decline in SBP and DBP, also reported in studies of older adults [22], may have made MVLTPA-related differences harder to detect. Finally, incident high blood pressure was defined using measurements from a single annual visit rather than a more detailed assessment, potentially introducing outcome misclassification and attenuating associations with MVLTPA [23]. Studies in younger adults with lower antihypertensive medication use and more frequent blood pressure measurements are warranted to clarify the relationship between habitual physical activity and blood pressure across the lifespan.

Despite the lack of clear differences in measured blood pressure, adequate MVLTPA was associated with a 17% lower risk of antihypertensive drug initiation, with a modest inverse linear dose-response association between MVLTPA and drug initiation. This is plausible because treatment initiation reflects clinical decisions based on overall cardiovascular risk [1]. MVLTPA-related improvements in adiposity may also reduce cardiovascular risk and, consequently, treatment requirements [24]. Exercise may also improve blood pressure control beyond isolated office measurements, as shown by reductions in ambulatory blood pressure [25], potentially reducing the need for antihypertensive medication. Higher LTPA has been previously associated with lower antihypertensive drug initiation [8] and randomized evidence also supports a role for exercise in reducing antihypertensive medication requirements among treated patients [26]. The association with antihypertensive drug initiation tended to be stronger among women, in agreement with a recent cohort study [27] and our previous findings of stronger MVLTPA associations with better adiposity among women [24]. Sex-related differences in physical activity patterns and their associations with cardiovascular health [28,29], together with potential differences in vascular responses to exercise [30] may contribute to this pattern, although we should be cautious with these mechanisms given the inconclusive interaction test by sex (*p*-value = 0.058).

This study is subject to several limitations. First, its observational design does not allow causal conclusions regarding the effect of MVLTPA on blood pressure, and the observed associations may be partly explained by residual confounding despite adjustment for multiple covariates. Reverse causation cannot be excluded: lower MVLTPA in older adults may partly reflect underlying frailty, mobility limitations, or early subclinical disease, which could themselves influence blood pressure. Our findings complement trial-based evidence by examining habitual MVLTPA over longer follow-up in an older, high-risk population. Future interventional studies, or triangulation with complementary epidemiological approaches such as Mendelian randomization [15,31], are warranted. Second, physical activity was assessed using a self-reported questionnaire, which may be subject to recall bias and measurement error. However, the use of cumulative averages of repeated measurements likely reduced random variability and better captured long-term exposure. Third, outcome assessment and treatment-related factors may have influenced our findings. The high prevalence of hypertension and use of antihypertensive medication may have obscured associations between MVLTPA and measured blood pressure. Furthermore, defining incident high blood pressure using annual measurements may have introduced outcome misclassification, while antihypertensive drug initiation depends on clinical decisions influenced by factors beyond blood pressure levels. Fourth, our longitudinal database combined repeated measurements from participants observed at different ages, without continuous follow-up across the entire 60-80-year age range. Consequently, trajectory estimates at different ages may be informed by different subsets of participants. We addressed this issue by 1) using mixed-effects spline models using all available repeated measurements; 2) restricting trajectory analyses to participants with at least two measurements; 3) applying inverse probability weighting in Kaplan-Meier analyses to address missingness; and 4) using age-as-time-scale Cox models treating loss to follow-up as right censoring. Nevertheless, missing data and informative loss to follow-up could still bias our estimates, particularly if missingness depends on unobserved factors. Studies with more complete longitudinal follow-up are needed to confirm these findings. Finally, the study population consisted of older adults at high cardiovascular risk from a Mediterranean setting, which may limit the generalizability of the findings to younger populations, lower-risk individuals, or non-Mediterranean populations.

## CONCLUSION

In this cohort of older adults at high cardiovascular risk, habitual MVLTPA was not associated with clinically meaningful differences in age-related SBP or DBP decline or with a lower risk of incident high blood pressure. However, adequate MVLTPA was associated with a 17% lower risk of antihypertensive medication initiation, with a modest inverse linear dose-response association and a potentially stronger association in women. By examining habitual MVLTPA over longer follow-up, our study complements evidence from exercise trials and highlights potential benefits for cardiovascular risk management beyond measured blood pressure. These findings support promoting regular physical activity among older adults at high cardiovascular risk, while further studies in younger, less medicated populations, incorporating repeated and out-of-office blood pressure measurements, are needed to clarify these associations and their clinical implications.

## DECLARATIONS

### Ethics approval and consent to participate

This study was conducted in accordance with the principles of the Declaration of Helsinki. The protocol was approved by the Ethics Committee of Parc de Salut Mar (reference 2024/11670/I; November 18, 2024). All participants provided written informed consent before enrolment in the PREDIMED study.

### Consent for publication

Not applicable.

### Availability of data and materials

The individual-level PREDIMED dataset cannot be made publicly available due to national data protection legislation, ethical restrictions, and the absence of explicit participant consent for open data sharing. Access to the data for bona fide collaborative research may be granted to qualified investigators upon reasonable request and approval by the PREDIMED Steering Committee. Requests should be addressed to. The code used for constructing the database, managing the data, and performing the statistical analyses is openly available at: https://github.com/alvarohernaez/MVLTPA_cardiometab_PREDIMED.

### Authors’ contributions

Conceptualization: Á.H. Methodology: E.F., Á.H.. Formal analysis: E.F., Á.H. Investigation: A.G., J.S.-S., R.E., E.R., E.G.-G., M.Fiol, J.L., J.T., L.S.-M., X.P., M.Fitó.

Data curation: Á.H. Writing-original draft: E.F. Writing-review and editing: M.H.H., A.G., A.C.-V., E.T., J.S.-S., R.E., E.R., E.G.-G., M.Fiol, J.L., J.T., L.S.-M., X.P., M.R.-C.,

H.V.-L., M.Fitó, Á.H. Funding acquisition: Á.H. Supervision: M.H.H., Á.H. All authors read and approved the final manuscript.

## Acknowledgements

We gratefully acknowledge all PREDIMED participants, investigators, and study personnel involved in participant recruitment, follow-up, data collection, and study coordination. CIBER de Enfermedades Cardiovasculares (CIBERCV) and CIBER de Fisiopatología de la Obesidad y Nutrición (CIBEROBN) are initiatives of Instituto de Salud Carlos III (Madrid, Spain) and are financed by the European Regional Development Fund.

## Supplementary information

Additional file 1 (.docx): Supplementary Tables 1 (Number of blood pressure observations by age) and Supplementary Figure 1 (Between-group differences in predicted values of systolic and diastolic blood pressure).

