## Supplemental Materials for "Moderate-to-vigorous physical activity, longitudinal blood pressure trajectories, and hypertension-related outcomes in older adults at high cardiovascular risk: a cohort study"

### SUPPLEMENTARY TABLES

**Supplementary Table 1.** Number of SBP and DBP observations available at each chronological age between 60 and 80 years.

| Age | SBP measures | DBP measures |
|---|---|---|
| 60 | 340 | 340 |
| 61 | 596 | 596 |
| 62 | 844 | 844 |
| 63 | 1,064 | 1,064 |
| 64 | 1,215 | 1,215 |
| 65 | 1,340 | 1,340 |
| 66 | 1,392 | 1,392 |
| 67 | 1,442 | 1,442 |
| 68 | 1,406 | 1,406 |
| 69 | 1,392 | 1,392 |
| 70 | 1,371 | 1,371 |
| 71 | 1,349 | 1,349 |
| 72 | 1,427 | 1,427 |
| 73 | 1,357 | 1,357 |
| 74 | 1,227 | 1,227 |
| 75 | 1,146 | 1,146 |
| 76 | 1,060 | 1,060 |
| 77 | 911 | 911 |
| 78 | 824 | 824 |
| 79 | 660 | 660 |
| 80 | 525 | 525 |

**SUPPLEMENTAL FIGURES**

**Supplementary Figure 1**. Between-group differences in systolic blood pressure (SBP, panel A) and diastolic blood pressure (DBP, panel B) between participants with adequate and insufficient MVLTPA across age groups. Results are presented for the overall population, stratified by sex, and among participants with hypertension at baseline, stratified by sex. Colored squares represent the following subgroups: black: men with hypertension at baseline; magenta: women with hypertension at baseline, blue: all participants with hypertension at baseline, green: men (overall population), dark red: women (overall population). orange: all participants (overall population). Values represent between-group differences in predicted SBP and DBP (mmHg), with corresponding 95% confidence intervals. The vertical dashed line indicates no difference between MVLTPA groups.

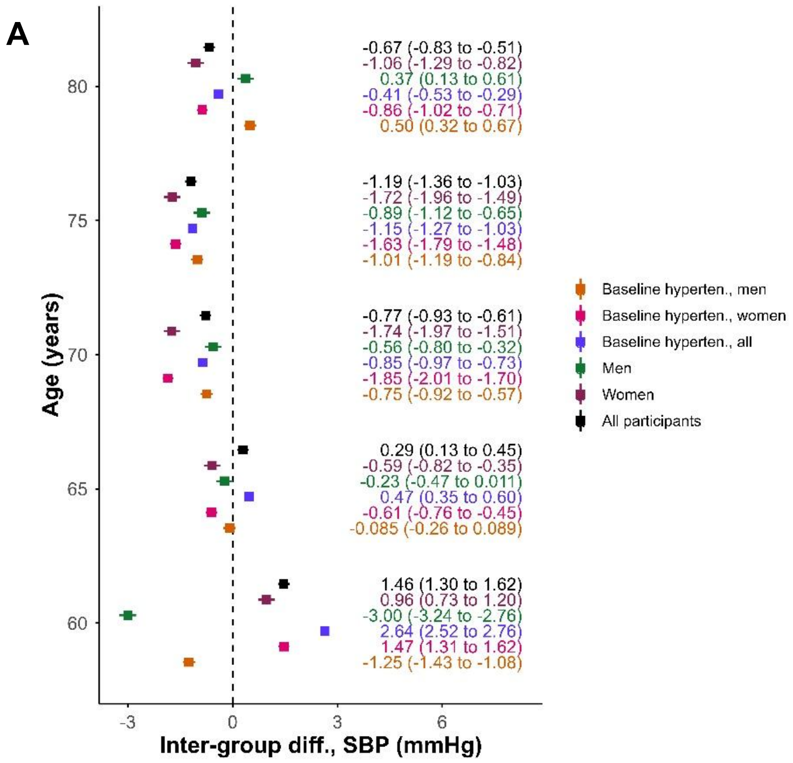

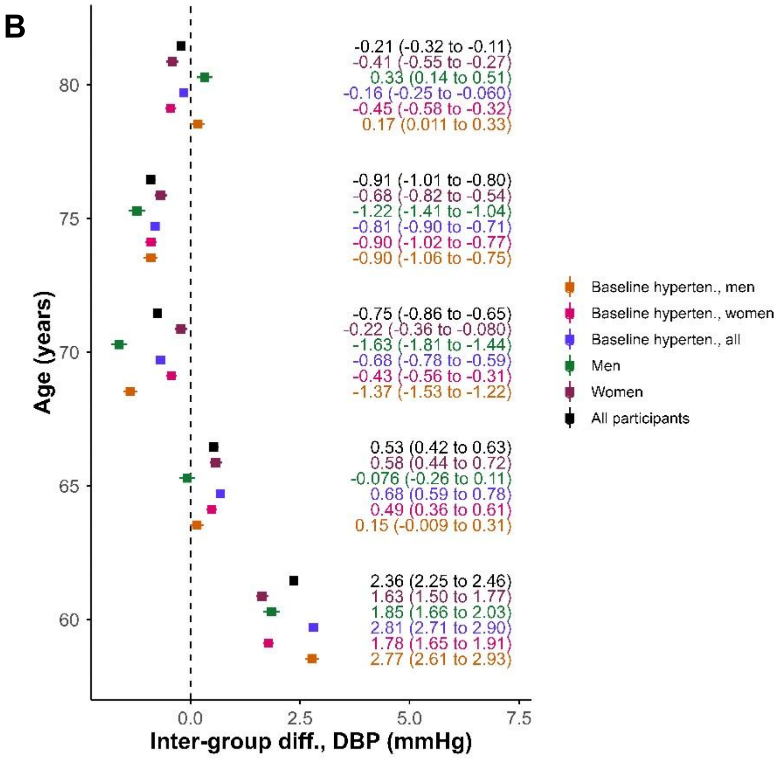
B
Age (years)
Inter-group diff., DBP (mmHg)
-0.21 (-0.32 to -0.11)
-0.41 (-0.55 to -0.27)
0.33 (0.14 to 0.51)
-0.16 (-0.25 to -0.060)
-0.45 (-0.58 to -0.32)
0.17 (0.011 to 0.33)
-0.91 (-1.01 to -0.80)
-0.68 (-0.82 to -0.54)
-1.22 (-1.41 to -1.04)
-0.81 (-0.90 to -0.71)
-0.90 (-1.02 to -0.77)
-0.90 (-1.06 to -0.75)
-0.75 (-0.86 to -0.65)
-0.22 (-0.36 to -0.080)
-1.63 (-1.81 to -1.44)
-0.68 (-0.78 to -0.59)
-0.43 (-0.56 to -0.31)
-1.37 (-1.53 to -1.22)
0.53 (0.42 to 0.63)
0.58 (0.44 to 0.72)
-0.076 (-0.26 to 0.11)
0.68 (0.59 to 0.78)
0.49 (0.36 to 0.61)
0.15 (-0.009 to 0.31)
2.36 (2.25 to 2.46)
1.63 (1.50 to 1.77)
1.85 (1.66 to 2.03)
2.81 (2.71 to 2.90)
1.78 (1.65 to 1.91)
2.77 (2.61 to 2.93)
Baseline hyperten., men
Baseline hyperten., women
Baseline hyperten., all
Men
Women
All participants
